# Evidence from imaging resilience genetics for a protective mechanism against schizophrenia in the ventral visual pathway

**DOI:** 10.1101/2021.07.23.21261045

**Authors:** Meike D. Hettwer, Thomas M. Lancaster, Eva Raspor, Peter K. Hahn, Nina Roth Mota, Wolf Singer, Andreas Reif, David E.J. Linden, Robert A. Bittner

## Abstract

**Introduction:** Illuminating neurobiological mechanisms underlying the protective effect of recently discovered common genetic resilience variants for schizophrenia is crucial for more effective prevention efforts. Current models implicate adaptive neuroplastic changes in the visual system and their pro-cognitive effects as a schizophrenia resilience mechanism. We investigated whether common genetic resilience variants might affect brain structure in similar neural circuits.

**Method:** Using structural magnetic resonance imaging, we measured the impact of an established schizophrenia polygenic resilience score (PRS_Resilience_) on cortical volume, thickness, and surface area in 101 healthy subjects and in a replication sample of 33,224 healthy subjects (UK Biobank).

**Finding:** We observed a significant positive whole-brain correlation between PRS_Resilience_ and cortical volume in the right fusiform gyrus (FFG) (r=0.35; p=.0004). Post-hoc analyses in this cluster revealed an impact of PRS_Resilience_ on cortical surface area. The replication sample showed a positive correlation between PRS_Resilience_ and global cortical volume and surface area in the left FFG.

**Conclusion:** Our findings represent the first evidence of a neurobiological correlate of a genetic resilience factor for schizophrenia. They support the view that schizophrenia resilience emerges from strengthening neural circuits in the ventral visual pathway and an increased capacity for the disambiguation of social and non-social visual information. This may aid psychosocial functioning, ameliorate the detrimental effects of subtle perceptual and cognitive disturbances in at-risk individuals, and facilitate coping with the cognitive and psychosocial consequences of stressors. Our results thus provide a novel link between visual cognition, the vulnerability-stress concept and schizophrenia resilience models.

## INTRODUCTION

The effective prevention of schizophrenia depends on a clear understanding of both risk mechanisms and mechanisms of resilience, which confer protection from this debilitating disorder. The emerging field of resilience research promises to provide novel pathways toward improved treatment and prevention strategies for psychiatric disorders.^1–4^

Resilience describes the phenomenon that many people with considerable exposure to risk factors for mental disorders retain good mental health.^5–7^ Importantly, current concepts regard resilience as a dynamic process facilitating adjustments to potentially disabling stressors, rather than as a trait or stable personality profile.^3, 6–8^ Nevertheless, there is clear evidence that resilience factors which contribute to resilience capacity^5, 6^ may well be genetically determined.^5, 9–15^

Resilience research in schizophrenia needs to acknowledge its complex, decade-spanning pathophysiological trajectory prominently involving neurodevelopmental disturbances^16–18^ and psychosocial stressors.^19, 20^ Moreover, schizophrenia is a disorder of information processing^21, 22^ featuring wide-ranging and pervasive perceptual and cognitive impairments.^23^ Schizophrenia resilience mechanisms might thus act over prolonged periods at multiple premorbid stages and should likely affect cognition. The high heritability of schizophrenia^24^ puts a particular emphasis on genetic mediators of resilience.

Concerted efforts have discovered common and rare genetic risk variants for schizophrenia.^25–27^ The additive effects of common variants have been captured by polygenic risk scores.^28^ Recently, first successful attempts at elucidating the genetic architecture of resilience^29^ have discovered single nucleotide polymorphisms (SNPs) moderating the penetrance of established common genetic risk factors.^30^ Resilience variants were identified by contrasting unaffected and affected individuals at an equally elevated polygenic risk^25^ to reveal residual genetic variation associated with resilience in high-risk but unaffected individuals. Schizophrenia risk variants were excluded to perform a genome-wide association study (GWAS) of resilience that detects effects on caseness, which are conditionally independent from risk variants. Thus, the identified resilience variants are not simply protective variants, i.e. the alternate alleles at risk loci. Rather, they are independent from and orthogonal to risk variants, addressing the concern that resilience must not simply be defined as the flip side of vulnerability.^5, 6^ Mirroring the concept of polygenic risk scores, the additive effects of these variants have been captured in the first polygenic resilience score (PRS*_Resilience_*) for schizophrenia,^29^ which reflects an individual’s genetic resistance to illness manifestation. This novel approach opens valuable opportunities to investigate how heritable mediators of resilience to schizophrenia influence neural systems to exert their protective effect.

Evidence for non-genetic schizophrenia resilience factors that could inform this research remains scarce. Presently, three putative protective mechanisms are most prominently discussed.^31, 32^ First, several pre- and perinatal factors appear to be protective against schizophrenia by increasing resilience to pregnancy-related and obstetric complications.^32^ Second, specific non-neurological auto-immune disorders, rheumatoid arthritis and ankylosing spondylitis, appear to be associated with a lower illness risk.^33^ Third, several resilience models emphasize the importance of the visual system.^31, 34, 35^ The latter are partly based on studies suggesting a reduced risk for schizophrenia in people with congenital/early (C/E) blindness,^33, 35–38^ which is neither observed in other forms of sensory loss, including C/E deafblindness and blindness acquired later in life,^35, 39–41^ nor for other mental disorders.^42–46^ Rather than to blindness per se, this has been attributed to adaptive reorganization of the visual system triggered by C/E blindness^47, 48^ and concurrent improvements in cognitive functions impaired in schizophrenia.^35, 40^ C/E blind individuals, but not late blind individuals show increased gray matter in parts of the inferior occipital, fusiform and lingual gyrus^49, 50^ as well as functional reorganization of both the ventral and dorsal visual pathway.^51, 52^

A useful framework for the interpretation of the postulated protective effect of C/E blindness is offered by the predictive coding theory of brain function.^53, 54^ Converging evidence indicates that key clinical and cognitive symptoms of schizophrenia arise from a decreased precision and stability of internal high-level priors relative to sensory information.^31, 55^ It has been argued that adaptive reorganization occurring in C/E blindness – and with it considerably greater reliance on context extracted from other sensory modalities – should improve the precision and stability of high-level and supramodal priors.^31^ The resulting greater primacy of priors relative to sensory data may facilitate resilience to schizophrenia.^31^

Importantly, current epidemiological evidence remains inconclusive due to the low base rates of both disorders.^56, 57^ Yet, while there is presently no consensus regarding the proposed role of C/E blindness as a resilience factor for schizophrenia,^31, 34, 35, 38, 40^ these models provide predictions that are testable using neuroimaging. Specifically, current concepts raise the question, whether similar but more subtle changes of the visual system exerting a weaker protective effect could also occur as a form of genetically mediated natural variation in sighted individuals.

A plethora of schizophrenia risk factors have been identified.^16, 32^ Similarly, multiple neurobiological pathways promoting resilience to the disorder would have to be expected. An involvement of the visual system in one of these resilience mechanisms would be conceivable given the prominent visual perceptual impairments featured in schizophrenia. These encompass basic deficits in visual acuity and contrast sensitivity^58–60^ and subsequent disturbances of perceptual organization including figure-ground segregation, coherent motion detection, contour integration, and perceptual closure.^61–66^ They further perturb higher-level visual processes, particularly object recognition,^61, 65, 67^ higher-order cognitive domains including working memory as well as social cognition,^62, 67–72^ and contribute to poor functional outcome.^73–75^ Abnormalities in both the dorsal and ventral visual pathway have been implicated.^61, 65, 76, 77^ Notably, visual dysfunction is among the strongest predictors of transition to full-blown illness in high-risk individuals^78, 79^ – more predictive than any other sensory anomalies and uniquely so for schizophrenia among mental disorders.^79^ Searching for resilience-promoting mechanisms within the visual system partly builds upon a successful strategy for risk research in neuropsychiatric disorders. Genetic studies indicate that rare but highly penetrant risk factors provide information about the neurobiological consequences of common genetic risk factors, which despite their lower penetrance tend to affect the same pathophysiological pathways.^80, 81^ C/E blindness as one potential, rare resilience factor with a profound impact on the brain could thus point to similar but less penetrant mechanisms of common resilience factors. Accordingly, SNPs conferring resilience to schizophrenia might exert their protective influence partly by affecting neuroplasticity in the visual system. To test this hypothesis and to investigate brain morphological correlates of genetic resilience to schizophrenia, we conducted an imaging genetics study using structural magnetic resonance imaging (sMRI) in healthy participants.

## METHODS AND MATERIALS

### Participants

All participants gave their written informed consent to participate in the study in accordance with the study protocol approved by the ethical review board of the Faculty of Medicine at Goethe University. The experimental procedures were conducted in conformity with the approved guidelines and the Declaration of Helsinki. We obtained structural MRI and genetic data from 105 right-handed subjects with normal or corrected to normal vision and no family or personal history of psychiatric disorders, according to the German Version of the SCID-I questionnaire for DSM-IV.^82^ Handedness was measured with the Edinburgh Handedness Inventory^83^ and IQ was determined using the MWT-B.^84^ We excluded four subjects due to excessive head motion during scanning (n=1) or substantial reconstruction errors during MRI processing (n=3), resulting in 101 subjects for the analysis (55 female; Mean age: 26.3 ± 4.71 years).

### Genotyping

We collected venous blood samples for DNA extraction. Genotyping was performed using custom Illumina HumanCoreExome-24 BeadChip genotyping arrays, which contain 570,038 genetic variants (Illumina, Inc., San Diego, CA). The Rapid Imputation and Computational Pipeline (RICOPILI^85^) was used for quality control, principal component analysis and imputation. Individuals were excluded for ambiguous sex, cryptic relatedness up to third degree relatives by identity of descent, genotyping completeness < 99 %, and non-European ethnicity admixture. SNPs were excluded where the minor allele frequency was < 1 %, if the call rate was < 99 % or if the χ^2^-test for Hardy-Weinberg Equilibrium had a p-value < 1 e-06. Before imputation, we assessed genetic homogeneity in each sample using multi-dimensional scaling (MDS) analysis. We excluded ancestry outliers through visual inspection of the first two components.

### Calculation of polygenic resilience scores

We calculated schizophrenia PRS*_Resilience_* based on resilience loci identified by Hess et al.,^29^ who included 3786 high-risk resilient individuals and 18,619 patients at equal polygenic risk in their initial discovery sample. Calculations of polygenic scores were performed according to previously described protocols with PRSice software v. 2.2.8.^86, 87^ Following the established PGC protocol,^25^ polygenic scores were clumped using linkage disequilibrium (LD) and distance thresholds of r^2^ = 0.1, within a 500 kb window. The major histocompatibility complex (MHC) region was excluded due to high LD.^25^ We calculated PRS*_Resilience_* based on SNPs thresholded at *p* < 0.05 because, among nominally significant PRS*_Resilience_* reported, SNPs included at this threshold were shown to explain the most variance.^29^

### Acquisition and analysis of MRI data

We acquired structural MRI data on a 3T Trio MR-scanner (Siemens, Erlangen, Germany) using a high-resolution T1-weighted Modified Driven Equilibrium Fourier Transform (MDEFT) sequence^88^ (voxel size: 1 x 1 x 1 mm³; TR = 7.92; TE = 2.4; TI = 910 ms; FOV = 256 x 256 mm^2^; number of slices per volume = 176; flip angle = 15°, slice thickness = 1 mm). We used FreeSurfer (version 6.0.1; http://surfer.nmr.mgh.harvard.edu) for semi-automated preprocessing and surface reconstruction, using bias-corrected^89, 90^ T1-weighted anatomical scans as the input (see Supplementary Materials). Measures of cortical thickness, cortical volume, and surface area were computed using FreeSurfer.^91, 92^ Surface maps were smoothed using a surface-based 10-mm full-width-half-maximum smoothing kernel.

### Statistical group analysis

Three general linear models (GLMs) were applied at each vertex to estimate the relationship between PRS*_Resilience_* and cortical volume, thickness, and surface area. Total intracranial volume (ICV) and age were included as covariates in partial correlation analyses. PRS*_Resilience_*, age and ICV were demeaned to allow for better model fit. To correct for multiple comparisons, we used surface-based cluster-size exclusion as implemented in FreeSurfer.^93^ We applied an initial cluster-forming threshold (CFT) of *p* < .001 and performed Monte Carlo simulations (10,000 iterations; cluster-wise *p* < .05, adjusted for testing both hemispheres separately, i.e. *p* < .025). These statistical parameters have been recommended to protect from type 1 errors.^94^

### UK Biobank (UKBB) replication analysis

To test the replicability of our findings, we performed additional analyses on GWAS summary statistics (https://open.win.ox.ac.uk/ukbiobank/big40/pheweb33k/phenotypes/) from a UKBB general population sample (n = 33,224).^95^ For this sample, vertex-wise data is not publicly available, precluding a whole-brain analysis. Rather, region of interest (ROI) based data is available in the standardized form of 33 Desikan-Killiany (DK) parcels per hemisphere (66 total), representing almost the entirety of the cerebral cortex.^96^ We used the well-established ‘gtx’ method^97–100^ (see Supplementary Methods) to test the association between PRS*_Resilience_* and cortical volume, thickness, and surface area in a ROI based manner. In keeping with the strictly confirmatory nature of this analysis, we focused on ROIs that best represent the location of clusters observed in the vertex-wise analysis in the discovery sample. Additionally, we compared effect sizes of these ROIs to the distribution of effect sizes of all ROIs, averaged across hemispheres, via z-tests.

## RESULTS

### Discovery sample

In our discovery sample, we observed a positive whole-brain correlation between PRS*_Resilience_* and cortical volume in the right fusiform gyrus (FFG) after correction for multiple comparisons (MNI: X = 33.7; Y = -51.4, Z = -16.4; cluster size: 338.1 mm^2^; *r* = 0.35; cluster-wise *p* = .0004; Figure 1A). Comparison with the neuroanatomical literature^101^ and probabilistic neuroimaging atlases of the ventral temporal cortex (VTC)^102–104^ indicated a position of this cluster on top of the mid-fusiform sulcus (MFS; see Supplementary Figure S1) and a partial overlap with the fusiform face area (FFA).^102–104^ Extracting the corresponding values from the FFG cluster revealed that PRS*_Resilience_* was significantly correlated with FFG surface area (*r* = .35, *p* < .001; Figure 1B) but not with cortical thickness (*r* = .14, *p* = .17; Figure 1C). We further observed an impact of PRS*_Resilience_* on left FFG volume at the initial CFT (p < .001), which, however, did not survive multiple comparisons correction (cluster-wise p > .05; Supplementary Figure S2). There was no significant correlation between PRS*_Resilience_* and either surface area or cortical thickness at the whole-brain level.

**Figure 1.**
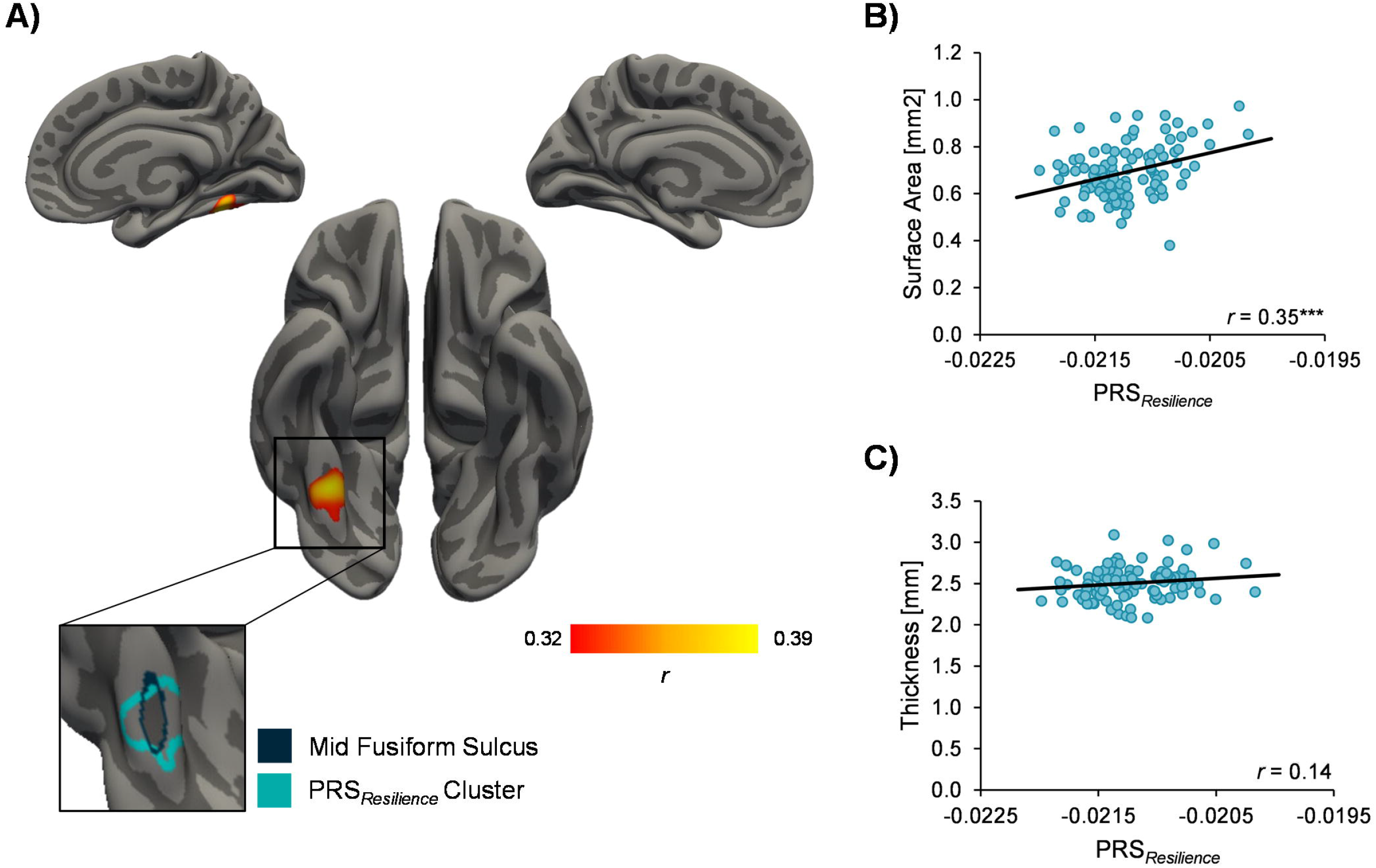
Whole-brain correlation between schizophrenia polygenic resilience scores (PRS*_Resilience_*) and cortical volume (cluster-wise p < .05 corrected) (A). Surface area (B) and cortical thickness (C) values in the right fusiform gyrus plotted against PRS*_Resilience_*.

### UKBB replication sample

Because the findings in our discovery sample hinted at a bilateral effect in the FFG, our replication analysis focused on left and right FFG parcels^96^ (Figure 2A). In the UKBB sample, we observed a significant effect of PRS*_Resilience_* on cortical volume (*p* = .006) and surface area (*p* = .026) in the left FFG but not in the right FFG (*p_volume_* = .208; *p_surface area_ =* .343; Figure 2B). PRS*_Resilience_* did not impact cortical thickness. Averaged across hemispheres, FFG effects of PRS*_Resilience_* on surface area (z = 2.5, *p* = .012) and cortical volume (z = 2.09, *p* = .036) were significantly higher than for all other ROIs (Figure 2C).

**Figure 2.**
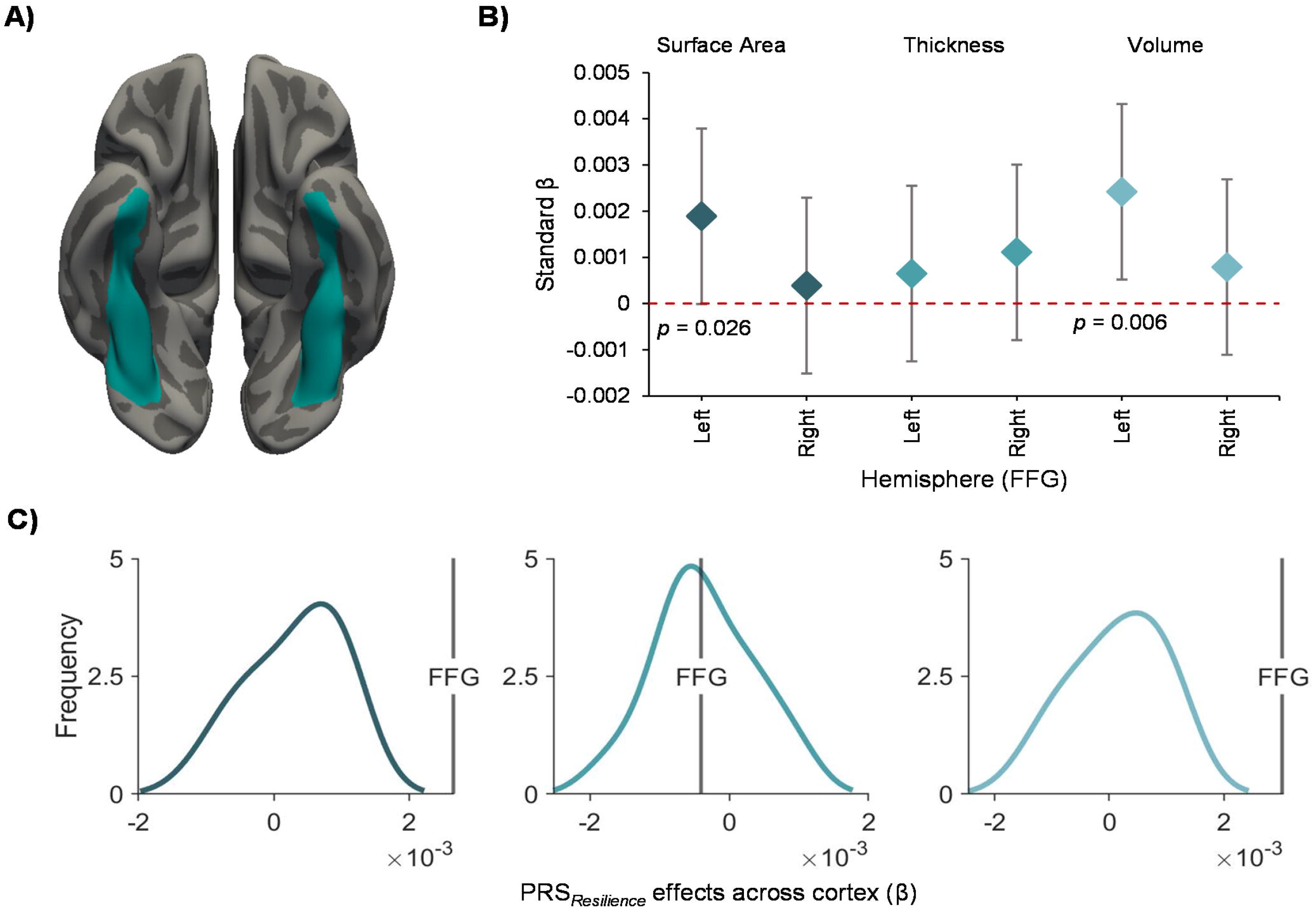
Effects of schizophrenia polygenic resilience scores (PRS*_Resilience_*) on fusiform gyrus (FFG) anatomical metrics in the UK Biobank sample. A) Desikan- Killiany^96^ (DK) FFG parcels. B) Significant PRS*_Resilience_* effects for cortical volume and surface area in the left FFG (error bars: 95% confidence intervals). C) PRS*_Resilience_* effects on FFG metrics averaged across hemispheres compared to the distribution of effect sizes (betas) for the other 32 DK parcels, also averaged for each anatomical region across hemispheres.

## DISCUSSION

We investigated the neuroanatomical correlates of common genetic variants associated with resilience to schizophrenia using sMRI. In the discovery sample, we observed a positive correlation between PRS*_Resilience_* and cortical volume in the right FFG, a key component of the ventral visual pathway. In the replication sample, we observed a correlation between global left FFG volume and PRS*_Resilience_*, which mirrors the cluster in the left FFG in the discovery sample that did not survive multiple comparisons correction. Involvement of the ventral visual pathway therefore appears not to be specific to one hemisphere. This is in line with evidence that both hemispheres – while independently analyzing half of the visual scene^105–107^ – combine their resources to jointly process objects at the fovea.^107^ Resilience to schizophrenia thus appears to result from a general increase in ventral visual pathway processing resources. Our findings provide the first direct evidence for a schizophrenia resilience mechanism involving the visual system in line with existing resilience models.

The particularly prominent effect in the FFG in comparison with all other bilateral ROIs indicates that, among cortical areas, the FFG plays a central role in promoting resilience to schizophrenia. Understanding the mechanisms underlying this protective effect requires a detailed assessment of the role of the FFG within the ventral visual pathway.

The ventral visual pathway is primarily involved in object recognition and categorization.^108–110^ Along this pathway, the FFG is an important relay between low level visual areas in the occipital lobe and areas in the infero-temporal cortex and para- hippocampal gyrus,^108, 109^ which are considered the top levels in the visual processing hierarchy. Importantly, all areas of the ventral stream from the primary visual cortex all the way up to the infero-temporal cortex are reciprocally connected via feed-forward and feed-back connections, communicating in parallel within various functional networks.^111, 112^ This connectivity profile, which also prominently includes interactions with occipital and parietal areas within the dorsal stream as well as prefrontal areas, is pivotal for shaping object and face recognition.^113–115^

The cluster observed in our discovery sample mapped directly onto the right MFS. This sulcus divides the FFG into a lateral and medial part, the two moieties differing in cyto- and receptor-architectonics^116, 117^ as well as their connectivity profiles within the VTC.^101, 118, 119^ The MFS further forms the central axis of multiple lateral-to-medial functional gradients across the VTC. These gradients relate to the eccentricity of the represented visual field, the size of objects, their animacy and domain specificity.^110, 120^ Described characteristics are indicative for the coexistence of functional networks devoted to the preferential processing of object categories.^110^ The increased volume of both moieties of the FFG as observed in our data might thus be indicative of a global enhancement of functions facilitating the parallel processing of complementary aspects of visual information.^107^ In the context of predictive coding,^53, 54^ this could imply greater resources for the disambiguation of sensory evidence by capitalizing on the priors stored in the functional architecture of the different processing streams. Notably, patients with schizophrenia are impaired in their ability to use stored knowledge for the interpretation of sensory evidence.^55, 121, 122^ Consequently, the embedding of sensory evidence in the context of a priori knowledge is disturbed. A larger FFG could compensate for disturbances that impede prior based evaluation of sensory information. This might also include an enhanced ability of the ventral stream to integrate input from the dorsal stream and from prefrontal areas, facilitating various facets of perceptual organisation during multiple stages of object processing, which is crucially impaired in schizophrenia.^65^ Such an interpretation is well in line with the notion that greater reliability of high-level priors might be protective against schizophrenia.^31^

Increased FFG volume may further contribute to resilience capacities through its involvement in interpersonal communication and social cognition.^120, 123–125^ Our cluster showed considerable overlap with the right FFA, the two subdivisions of which are located approximately at anterior and posterior ends of the MFS.^101, 110, 126^ The FFA is essential for the processing of face-related information including the decoding of facial components for affect discrimination^120, 123, 124^ and an important perceptual node of social cognitive networks.^125^ In schizophrenia, reduced FFG volume acts as a structural neural substrate of various perceptual and affective processing deficits associated with impaired social cognition.^67, 127–131^ FFG volume further contributes reliably to discriminative predictive patterns of social functioning and psychosis transition in high-risk individuals.^132, 133^ Importantly, perceptual deficits such as difficulties to decode complex facial configurations accompany and likely precede aberrant cognitive and affective processing in impaired social cognition.^57, 123, 127–129^ Our data suggest that genetically mediated resilience mechanisms involving the FFA might facilitate social perceptual processing capacities. This hypothesis is supported by reports of increased FFG volume in healthy subjects after social cognitive training.^137^ Furthermore, larger FFG volume in patients with schizophrenia appears to mediate the beneficial effects of cognitive training on general neurocognition and social cognition in particular.^138^ Together, these findings underscore that increased FFG volume is indeed associated with enhanced cognitive capabilities in multiple domains. They are also compatible with the role of the FFG as a central relay between low level visual and higher cognitive areas in support of a wide array of non-social and social cognitive processes.

A crucial role of social cognition for resilience to schizophrenia is conceivable, as it constitutes a major predictor of functional outcome^125^ and can compensate for the deleterious effects of neurocognitive deficits on daily life.^139–144^ Therefore, FFG related resilience mechanisms enhance cognitive functions which facilitate the embedding of individuals in their social environment, thus buffering against the effects of risk factors. Expanded computational resources in the ventral visual pathway should also facilitate coping with stressors resulting from sensory deficits. Typically, resilience models emphasize the ability to cope with stressors such as challenging life circumstances and physical illness.^3, 6, 145^ However, perturbed visual information processing in schizophrenia might in itself constitute a stressor. In addition to deficits in perceptual organization, characteristic disturbances comprise a lower threshold for sensory overload,^146^ visual distortions,^146, 147^ reduced predictability of changes in the environment and resulting uncertainty regarding adequate behavior^31^ as well as psychotic experiences.^75, 148^ These in turn have detrimental effects on adaptive behavior and interpersonal interactions, together constituting a fundamental stressor. Given the widespread deleterious impact of stress on cognition,^149, 150^ this might lead to a vicious cycle of increasing psychosocial stress, aberrant information processing and psychosis proneness.^20^ Through increased perceptual and cognitive capabilities, the resilience mechanisms implied by our data should reduce stress induced by subtle perceptual impairments and increase the chances of interrupting this vicious cycle. It remains to be clarified whether increased FFG resources promote a resilience mechanism that directly antagonizes potential cognitive disturbances or rather mitigates cognitive deficits by compensating for disease-related abnormalities. The latter would be consistent with the idea that, on the neurobiological level, resilience does not simply reverse pathophysiological processes but rather ameliorates the harmful consequences of stressors.^3^

Overall, our findings point towards a central role of visual cognition in schizophrenia resilience as both a stressor and a coping mechanism. Furthermore, our results provide a crucial link between genetics, the information processing disorder concept and the vulnerability-stress model.^22^ Interestingly, there is preliminary evidence for an involvement of the FFG and neighboring occipito-temporal areas in resilience against stress and adverse life events,^151^ trauma,^152^ and bipolar disorder,^153, 154^ suggesting a potential transdiagnostic relevance of visual cognition as a resilience mechanism.

Lastly, current findings create an interesting link to resilience models derived from C/E blindness research.^34^ They suggest that SNPs associated with resilience to schizophrenia can drive some of the neuroplastic changes in visual areas that are also observed in early reorganization following C/E blindness.^49, 50^ The convergence of such widely different biological mechanisms on a similar intermediate phenotype underscores the significance of the visual brain for schizophrenia resilience research. Importantly, this also raises the possibility that similar protective adaptations might be inducible through targeted interventions.^75^ Such interventions would likely have to occur at early stages of post-natal development when the visual system is susceptible to environmental influences.^155, 156^ This is suggested by the lack of a protective effect in late blindness,^35^ the early start and prolonged trajectory of abnormal neurodevelopment in schizophrenia^16^, as well as the early manifestation of visual dysfunction in at-risk populations.^78, 157–159^

Our findings provide a novel direction for schizophrenia resilience research by demonstrating that resilience to the disorder might arise from genetic influences that act on neural systems subserving elementary cognitive processes. However, the indirect nature of the current evidence is an important limitation of our study. The cognitive implications of our findings can so far only be inferred from existing evidence for an association of FFG volume and cognition and the crucial functional role of the FFG within the ventral visual pathway. Here, a direct investigation of links between cognition and PRS*_Resilience_* effects on FFG morphology was limited to publicly accessible cognitive measures in the UKBB (see Supplementary Material), which are not tailored toward schizophrenia research and our findings specifically. Furthermore, the current imaging genetics approach cannot provide evidence that individuals with higher PRS*_Resilience_* would indeed be more adaptable to the specific stressors implicated by our findings. These questions need to be addressed in future studies. Moreover, while our data suggest that structural neuroplastic alterations in the FFG contribute most prominently to schizophrenia resilience compared to other cortical areas, neuroimaging methods directly assessing brain function as well as structural and functional connectomics will be essential to elucidate the underlying mechanisms more completely. It is also highly likely that these methods will reveal additional, potentially unrelated resilience promoting neural circuits that sMRI is not sensitive for.

In conclusion, our study contributes to models of schizophrenia resilience by demonstrating an impact of genetic variants conferring protection from the disorder on local brain structure in the visual system. Improved cognitive and functional capacities that may result from these effects suggest a resilience mechanism linked to coping with both the cognitive and psychosocial sequelae of stressors. Future studies should investigate directly the individual contributions of the cognitive processes implicated by our findings to schizophrenia resilience in healthy and at-risk populations. Moreover, it will be imperative to investigate the interaction between genetic and environmental risk factors with resilience factors in order to fully realize the potential of the resilience paradigm for the discovery of novel course-modifying and preventive interventions for schizophrenia.

## Data Availability

All data needed to evaluate the conclusions in the paper are present in the paper and/or the Supplementary Materials. The data can be provided by Robert A. Bittner pending scientific review and a completed material transfer agreement. Requests for the data should be submitted to: Robert A. Bittner.

## ACKNOWLEDGEMENTS

The authors thank Christina Novak and Astrid Rehner for help with recruiting and data acquisition. We also thank Caroline Mann and Thorsten Kranz for advice regarding data processing and genetic analyses, respectively.

## AUTHOR CONTRIBUTIONS

Conceptualization: MDH, RAB

Methodology: MDH, TL, ER, NRM, RAB

Investigation: PKH, MDH, TL, RAB

Visualization: MDH

Supervision: RAB

Writing – original draft: MDH, RAB

Writing – review and editing: MDH, TL, WS, AR, DEJL, RAB

## AVAILABILITY OF DATA AND MATERIALS

The datasets generated during and/or analyzed during the current study are available from the corresponding author on reasonable request.

