## Supplementary Material for "Evidence from imaging resilience genetics for a protective mechanism against schizophrenia in the ventral visual pathway"

### Supplementary Methods

*Freesurfer image processing pipeline*

All anatomical magnetic resonance imaging (MRI) scans were processed in FreeSurfer (version 6.0.1; http://surfer.nmr.mgh.harvard.edu) using default parameters during recon-all and bias-corrected[1,2] T1 images as the input. The extensively validated procedures implemented in FreeSurfer are described in more detail elsewhere.[3–8] Briefly, the pipeline included motion correction, bias field correction, brain extraction with removal of cerebellum and brainstem, and spatial normalization. A triangular mesh covering the external white/grey matter boundary as determined by voxel intensity values was created. Deformation, expansion towards the pial (i.e. outer grey matter) surface, and topology correction fixing potential structural abnormalities was performed. Triangular cortical meshes for white and pial surfaces consisted of ~150.000 vertices per hemisphere. Surface reconstructions were manually checked for inaccuracies and corrected following the editing guidelines provided on the FreeSurferWiki (http://freesurfer.net/fswiki/Edits).

*UK Biobank replication analysis*

We investigated effects of $PRS_{Resilience}$ on brain structural parameters in the UKBB sample using the 'gtx' method,[9–12] which is equivalent to the "inverse variance weighted" approach in mendelian randomisation studies. However, in a polygenic score analysis there are no stringent inclusion criteria for genetic variants: we do not require the variants to be strongly associated with the disease and pleiotropic effects are allowed. All GWAS summary data was corrected for demographic and genetic confounds.[13] $PRS_{Resilience}$ were created using PRSice v1.25 risk profile software[14] with a stringent clumping procedure [clump.kb = 1000 kb, clump.r2 = 0.01] to remove correlated alleles. SNPs in the 1000 Genomes Project (phase 3) were used as reference data. We included SNPs thresholded at $p < 0.05$ from the schizophrenia resilience GWAS.[15] Variants within the MHC were removed from data (chr 6: 26,000–34,000 kb).

*Extended genetic analysis of FFG structure in UKBB*

We further sought to investigate the shared genetic etiology of FFG structure (volume, thickness, area, N = 33224) with cognitive ability (Fluid intelligence, N = 117,131; Digit span (maximum digits remembered, N = 36,884), and visual acuity (logarithm of the minimum angle of resolution (logMAR) for the left and right eye, N = 79,293). LD score regression[16] was first used to demonstrate significant heritability for all FFG image-derived phenotypes ($h^2 > 0.1$, $p < 0.001$ in cases), cognitive ($h^2 > 0.1$, $p < 0.001$) and visual acuity phenotypes ($h^2 > 0.028$, $p = 0.001$). We then proceeded to use LD score regression to estimate pairwise genetic correlations between FFG structure, cognitive and visual acuity phenotypes using LD scores derived from European 1000 genomes sample.

### Supplementary Results

*Behavioral results*

Visual acuity, fluid intelligence, and maximum digits remembered in the digit-span task did not show a significant genetic relationship with FFG volume, surface area, or thickness after correction for multiple comparisons (FDR 0.05; See Table S1).

**Table S1.** Genetic relationships between FFG morphology and behavioral parameters in the UKBB.

| | Hemisphere | Metric | rg | se | z | p | p(FDR) |
|---|---|---|---|---|---|---|---|
| Fluid intelligence | LH | Area | 0.0275 | 0.0423 | 0.6503 | 0.5155 | 0.7498 |
| Fluid intelligence | RH | Area | 0.0622 | 0.0415 | 1.4985 | 0.134 | 0.3573 |
| Fluid intelligence | LH | Volume | -0.0009 | 0.0466 | -0.0194 | 0.9846 | 0.9846 |
| Fluid intelligence | RH | Volume | 0.0493 | 0.0425 | 1.1600 | 0.2461 | 0.5625 |
| Maximum digits remembered | LH | Area | 0.079 | 0.0687 | 1.1498 | 0.2502 | 0.5004 |
| Maximum digits remembered | RH | Area | 0.1291 | 0.0722 | 1.7871 | 0.0739 | 0.2956 |
| Maximum digits remembered | LH | Volume | 0.0268 | 0.0784 | 0.3415 | 0.7327 | 0.9018 |
| Maximum digits remembered | RH | Volume | 0.0343 | 0.0745 | 0.4605 | 0.6452 | 0.8602 |
| Right logMAR | LH | Area | -0.197 | 0.0906 | -2.1746 | 0.0297 | 0.4752 |
| Right logMAR | RH | Area | -0.2121 | 0.1002 | -2.1178 | 0.0342 | 0.2736 |
| Right logMAR | LH | Volume | -0.1091 | 0.0977 | -1.1176 | 0.2637 | 0.4688 |
| Right logMAR | RH | Volume | -0.166 | 0.1011 | -1.6418 | 0.1006 | 0.3219 |
| Left logMAR | LH | Area | -0.2016 | 0.0969 | -2.0803 | 0.0375 | 0.2000 |
| Left logMAR | RH | Area | -0.0701 | 0.1032 | -0.6795 | 0.4968 | 0.7949 |
| Left logMAR | LH | Volume | -0.0319 | 0.1084 | -0.2944 | 0.7685 | 0.8782 |
| Left logMAR | RH | Volume | -0.0058 | 0.1101 | -0.0527 | 0.9579 | 1 |

FDR = False Discovery Rate using Benjamin Hochberg Correction.

**Supplementary Figures**

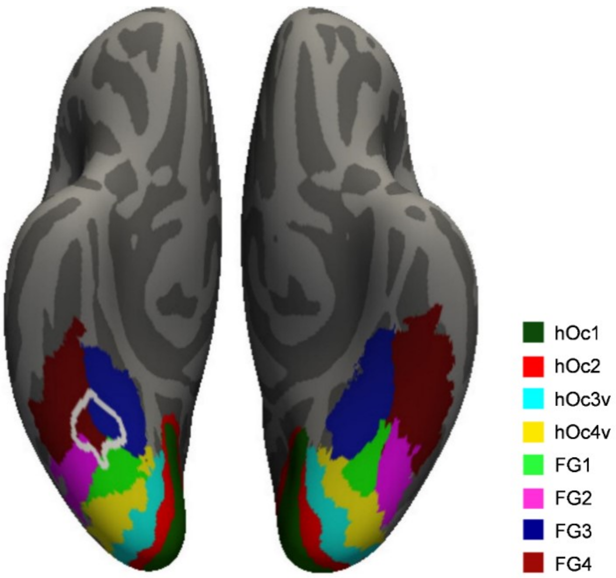


**Figure S1.**

Contextualization of fusiform gyrus cluster with a cytoarchitectonic atlas of the ventral visual stream. The cytoarchitectonic atlas published by Rosenke et al.[17] was rendered onto the inflated fsaverage surface in FreeSurfer. White outlines in the right hemisphere indicate the position of the fusiform gyrus cluster reflecting the impact of polygenic resilience scores on cortical volume described in the main manuscript. Occipital regions = hOc1-hOc4v; Fusiform gyrus sub-regions: FG1-FG4.

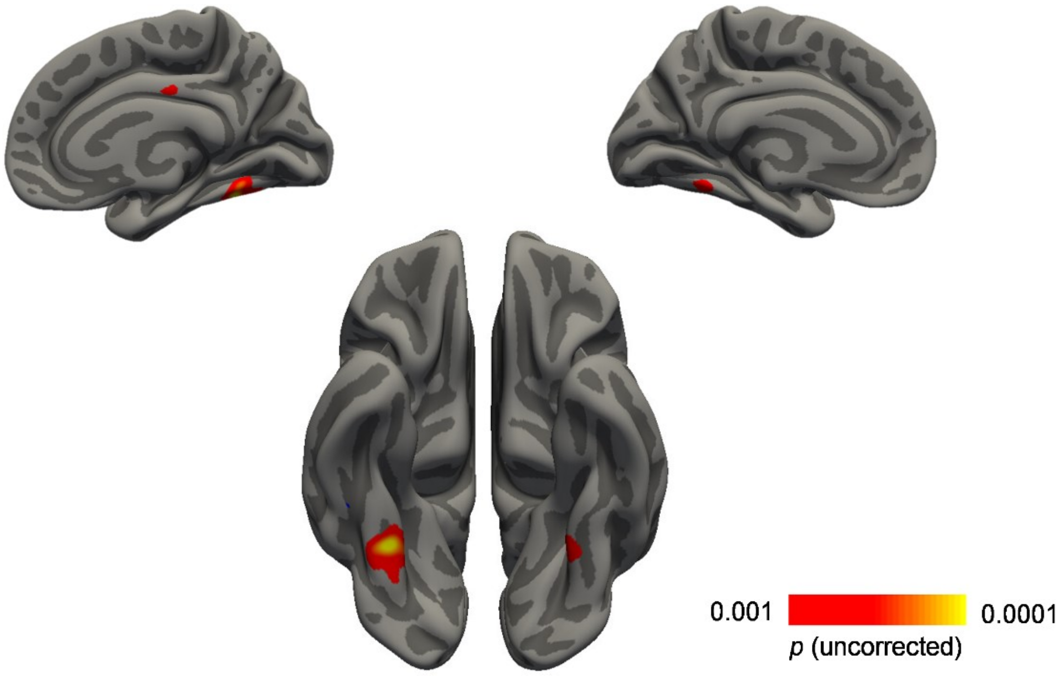


**Figure S2.**

Whole-brain correlation between $PRS_{Resilience}$ and cortical volume at cluster-forming threshold: $p < .001$. Results are shown for the discovery sample ($n = 101$) and are corrected for age and intracranial volume, but without correction for multiple comparisons using cluster thresholding. The bilateral effect on fusiform gyrus volume as observed at the level of vertex-wise thresholding did not survive multiple comparisons correction in the right hemisphere (see main manuscript).

17. Rosenke M, van Hoof R, van den Hurk J, Grill-Spector K, Goebel R. A

Probabilistic Functional Atlas of Human Occipito-Temporal Visual Cortex. *Cereb Cortex*. 2021;31(1):603-619. doi:10.1093/cercor/bhaa246
